# An interpretable Day-1 machine-learning model for COVID-19 mortality and severity prognosis: development in a Pakistani hospital cohort and external evaluation in Chinese and Italian populations

**DOI:** 10.64898/2026.10.01.26364553

**Authors:** Talha Jaffer, Shakir Aslam, Nida Javaid, Aasia Khaliq, Eesha Tariq, Tehmina Mustafa, Muhammad Adeel Zaffar, Shaper Mirza

## Abstract

Early risk stratification of COVID-19 patients at admission informs triage and resource allocation, particularly in low- and middle-income settings where intensive-care capacity is constrained. Most published models are not interpretable, rarely externally validated, and developed almost exclusively in high-income cohorts. We developed an interpretable random-forest classifier for three-class COVID-19 severity (Mild: not ventilated, survived; Severe: ventilated, survived; Fatal: died) using only data available within 24 hours of admission in 321 hospitalised PCR-confirmed COVID-19 patients in Pakistan. From 2,278 raw variables we retained 20 predictors by clinical screening and permutation-importance selection, with leakage-controlled multiple imputation by chained equations and SHapley Additive exPlanations (SHAP) for interpretation. A 15-feature variant was evaluated in two independent public cohorts: the Chinese iCTCF cohort (n = 894) and an Italian complete-blood-count cohort (n = 1,218). The development model achieved a macro F1 of 0.55, identifying the Fatal class most reliably (F1 0.67, recall 0.73) and Mild cases least well. SHAP identified a parsimonious set of admission-time predictors (oxygen saturation, lactate dehydrogenase, urea, C-reactive protein, age and radiographic pneumonia), including a raised urea-to-creatinine ratio in fatal cases most consistent with dehydration (median urea 80 vs 50 mg/dL, fatal vs mild; p < 0.001). Externally, the model retained useful discrimination in the Chinese cohort (mortality AUC 0.82 in 719 patients with recorded outcomes; severe-or-worse AUC 0.75; macro F1 0.50) but only modest discrimination in the Italian cohort (AUC 0.62, 95% CI 0.56-0.68), which we treat as a feature-availability stress test rather than a second validation. Cross-cohort SHAP rankings correlated strongly in China (Spearman ρ = 0.91) and moderately in Italy (ρ = 0.76), where most laboratory predictors were imputed and carried no information beyond the complete blood count; discrimination tracked how many predictors were actually measured. The admission-time signature converges with established markers of hypoperfusion and inflammation.

**Author summary:** When patients arrive at hospital with COVID-19, risk must be assessed immediately to direct limited resources to those in need, particularly where intensive-care capacity is scarce, as in Pakistan. Most existing computer-based tools come from developed countries, act as “black boxes”, and are rarely tested elsewhere. We built a model using first-day admission data from 321 patients at a hospital in Lahore, Pakistan. It identifies patients who later died best, and milder cases least well. The strongest markers were blood oxygen, lactate dehydrogenase, urea, C-reactive protein, age and chest-imaging evidence of pneumonia, together pointing to a coherent picture of pulmonary impairment, inflammation and circulatory stress. In fatal cases urea was markedly raised while creatinine rose only modestly, a pattern that most likely reflects dehydration. We then tested the same model on independent Chinese and Italian cohorts: it transferred well to China but lost accuracy in Italy, where only a basic blood count was available. The findings support an interpretable Day-1 prognostic tool whose useful range is set largely by which laboratory tests are available rather than by geography. Because these markers reflect general inflammation rather than anything unique to COVID-19, the approach may extend to other pneumonias.

## Introduction

Early risk stratification of hospitalised COVID-19 patients informs triage and resource allocation, particularly in low- and middle-income countries (LMICs) where intensive-care capacity is constrained [1]. Several machine-learning prognostic models have been proposed for this purpose, often reporting areas under the receiver-operating-characteristic curve (AUC) above 0.85 [2]. However, systematic reviews have repeatedly flagged these models at high risk of bias, citing data leakage, the use of features unavailable at admission, small or non-representative samples, and an almost universal absence of external validation [3,4]. Few are clinically interpretable in a way that lets clinicians see what the model is reacting to. This limits trust, slows adoption, and obscures whether predictions rely on coherent clinical signals or on cohort-specific artefacts [5,6].

Two gaps motivate this work. First, evidence from LMIC populations is limited: most models are developed and externally validated only within high-income cohorts, leaving their behaviour in resource-constrained settings unknown [3,7]. Second, external validation is usually reported only as a change in discrimination (a drop in Area Under the Curve [AUC]), without examining whether the model continues to rely on the same predictors in the new setting. Whether discrimination arises because the underlying population differs, or because the model’s required inputs are unavailable, has very different implications for deployment and revision [8,9].

We addressed both gaps. We developed an interpretable three-class severity model (Mild, Severe, Fatal) from a Pakistani hospital cohort using only Day-1 admission data, with SHapley Additive exPlanations (SHAP), which attribute each prediction to the contribution of its individual features, for interpretation [5,10]. We then externally validated the same model against two independent international cohorts: the Chinese iCTCF cohort (Wuhan) [11] and an Italian complete-blood-count cohort (San Raffaele, Milan [12]), which differ in geography, prevalence of severity and feature availability. Beyond discrimination, we use cross-cohort SHAP to test whether the model’s predictive signature is preserved across settings, separating population mismatch from input-availability limitations. We report the study in accordance with the TRIPOD+AI recommendations for prediction-model studies [8] and the STROBE statement for observational studies [13].

## Results

### Development cohort and model performance

Of 321 hospitalised PCR-confirmed patients, 81 (25.2%) were Mild, 132 (41.1%) Severe and 108 (33.6%) Fatal; 211 (65.7%) received mechanical ventilation. Baseline characteristics differed across severity classes (Table 1). Fatal patients were older (median age 60 vs 56 years in mild patients, p = 0.0083) and had lower SpO₂ (91.5% vs 94%, p < 0.001) but higher LDH (770 vs 490 U/L, p < 0.001), urea (80 vs 50 mg/dL, p < 0.001) and CRP (46.7 vs 18.1 mg/L, p = 0.016). Cardiovascular disease was more prevalent in fatal cases (33.3% vs 9.9%, p < 0.001), as were pneumonia infiltrates on admission imaging (72.2% in fatal vs 58.0% in mild cases, p < 0.001 across the three classes). Both imaging findings were non-monotonic across the severity classes, being least frequent in the Severe group (infiltrates 48.5%, ground-glass opacity 25.0%); this is consistent with the Severe class being defined by receipt of mechanical ventilation rather than by admission radiographic burden. Sex distribution and most other comorbidities did not differ significantly between classes.

**Table 1.** Baseline characteristics of the development cohort by severity class. All values are pre-imputation. Continuous variables are reported as median [interquartile range]; binary variables as n/N (%), where N is the number of patients in whom the variable was recorded. The n in each row label is the number of patients contributing to that row. p values are from Kruskal-Wallis tests (continuous) or chi-square tests (binary). Age was not recorded for 27 patients and sex for 4; laboratory tests were recorded in 29.6-75.4% of patients depending on the test. Missing values were multiply imputed for modelling (see Materials and Methods). Comorbidity categories are not mutually exclusive: 252 of 321 patients had at least one and individual patients had up to 5, so the counts in that block sum to more than the number of patients in each class.

| Characteristic | Mild (n=81) | Severe (n=132) | Fatal (n=108) | Total (n=321) | p |
| --- | --- | --- | --- | --- | --- |
| **Demographics** |  |  |  |  |  |
| Age, years, median [IQR] (n = 294) | 56 [45-61] | 58 [50-64] | 60 [55-72] | 58 [50-65] | 0.0083 |
| Male sex, n/N (%) | 49/79 (62.0) | 96/131 (73.3) | 73/107 (68.2) | 218/317 (68.8) | 0.23 |
| **Day-1 vital signs and laboratory values, median [IQR]** |  |  |  |  |  |
| SpO<sub>2</sub>, % (n = 242) | 94 [92-95] | 94 [92-96] | 91.5 [88-94] | 93 [90-95] | <0.001 |
| LDH, U/L (n = 166) | 490 [360-638] | 536 [385-767] | 770 [531-1019] | 590 [410-832] | <0.001 |
| Urea, mg/dL (n = 150) | 50 [40-68] | 50 [38-70] | 80 [51-150] | 61 [42-91] | <0.001 |
| CRP, mg/L (n = 198) | 18.1 [4.5-89] | 21.9 [8.9-62] | 46.7 [12.4-123] | 25.2 [8.4-93] | 0.016 |
| Creatinine, mg/dL (n = 179) | 0.9 [0.8-1.05] | 0.9 [0.8-1.25] | 1.1 [0.8-2.27] | 1.0 [0.8-1.3] | 0.11 |
| D-dimer, µg/mL (n = 117) | 1.6 [1.0-3.3] | 2.7 [1.1-6.9] | 3.3 [1.8-6.2] | 2.6 [1.2-5.6] | 0.040 |
| TLC, ×10<sup>3</sup>/µL (n = 214) | 12.8 [8.4-17.0] | 12.5 [9.0-15.8] | 14.2 [11.0-19.4] | 13.1 [9.4-18.1] | 0.084 |
| Haemoglobin, g/dL (n = 185) | 12.9 [11.4-14.0] | 12.8 [11.3-13.7] | 12.7 [11.3-14.2] | 12.8 [11.3-14.0] | 0.73 |
| Platelets, ×10<sup>3</sup>/µL (n = 179) | 253 [204-299] | 244 [183-291] | 233 [142-312] | 247 [182-304] | 0.71 |
| ALT, U/L (n = 98) | 60.5 [44.5-90.2] | 63.0 [34.2-84.2] | 47.5 [28.8-65.5] | 53.5 [33.2-78.8] | 0.14 |
| AST, U/L (n = 95) | 56 [40-82] | 67 [38-109] | 57 [34-96] | 58 [36-101] | 0.82 |
| **Comorbidities, n/N (%)** |  |  |  |  |  |
| Cardiovascular disease | 8/81 (9.9) | 32/132 (24.2) | 36/108 (33.3) | 76/321 (23.7) | <0.001 |
| Hypertension | 41/81 (50.6) | 80/132 (60.6) | 67/108 (62.0) | 188/321 (58.6) | 0.24 |
| Diabetes | 29/81 (35.8) | 65/132 (49.2) | 56/108 (51.9) | 150/321 (46.7) | 0.069 |
| Chronic kidney disease | 3/81 (3.7) | 10/132 (7.6) | 12/108 (11.1) | 25/321 (7.8) | 0.17 |
| Current smoking | 3/81 (3.7) | 11/132 (8.3) | 9/108 (8.3) | 23/321 (7.2) | 0.38 |
| **Imaging findings on admission, n/N (%)** |  |  |  |  |  |
| Pneumonia infiltrates | 47/81 (58.0) | 64/132 (48.5) | 78/108 (72.2) | 189/321 (58.9) | <0.001 |
| Ground-glass opacity | 37/81 (45.7) | 33/132 (25.0) | 46/108 (42.6) | 116/321 (36.1) | 0.002 |

In model development, the 20-feature subset and the full 32-feature model performed equivalently (cross-validated macro F1 0.55 ± 0.09 for both, a difference of 0.001 that is negligible relative to the between-fold spread), and both were clearly above the smaller subsets (Top 15: 0.52; Top 10: 0.50; Top 7: 0.49) (Table 2). The 20-feature subset was therefore retained for parsimony rather than on the basis of a measurable performance advantage. Hyperparameter tuning did not improve performance: the default-hyperparameter random forest (macro F1 0.55 ± 0.09, balanced accuracy 0.56 ± 0.09, accuracy 0.58) outperformed the tuned XGBoost (0.54), tuned random forest (0.52) and tuned gradient boosting (0.51), consistent with overfitting at this sample size [14].

**Table 2.** Feature-subset comparison from 10-fold cross-validation. The 20-feature subset and the full 32-feature set performed equivalently, and the 20-feature subset was retained as the primary model for parsimony.

| Feature subset | n features | Macro F1 | Balanced accuracy |
| --- | --- | --- | --- |
| Top 7 | 7 | 0.494 ± 0.111 | 0.499 ± 0.110 |
| Top 10 | 10 | 0.498 ± 0.084 | 0.509 ± 0.086 |
| Top 15 | 15 | 0.516 ± 0.100 | 0.531 ± 0.101 |
| **Top 20 (primary)** | **20** | **0.551 ± 0.086** | **0.560 ± 0.087** |
| All features | 32 | 0.550 ± 0.092 | 0.560 ± 0.091 |

The Fatal class was identified most reliably (F1 0.672; precision 0.622, recall 0.731), reflecting its distinctive biochemical profile, followed by Severe (F1 0.580) and Mild (F1 0.414). Most of the model’s discriminative power therefore lay in identifying Fatal outcomes; Mild cases were separated least well, reflecting their overlap with Severe disease on admission-time markers, a boundary that is itself defined by receipt of mechanical ventilation rather than by a purely physiological state (Table 3; Fig 1). SpO₂ was the dominant predictor (permutation importance 0.103, more than double the next feature)^1^, followed by LDH (0.047), age (0.046), D-dimer (0.037), creatinine (0.036), CRP (0.034), TLC (0.033), ALT (0.030), platelets (0.028) and urea (0.024).

**Fig 1.**
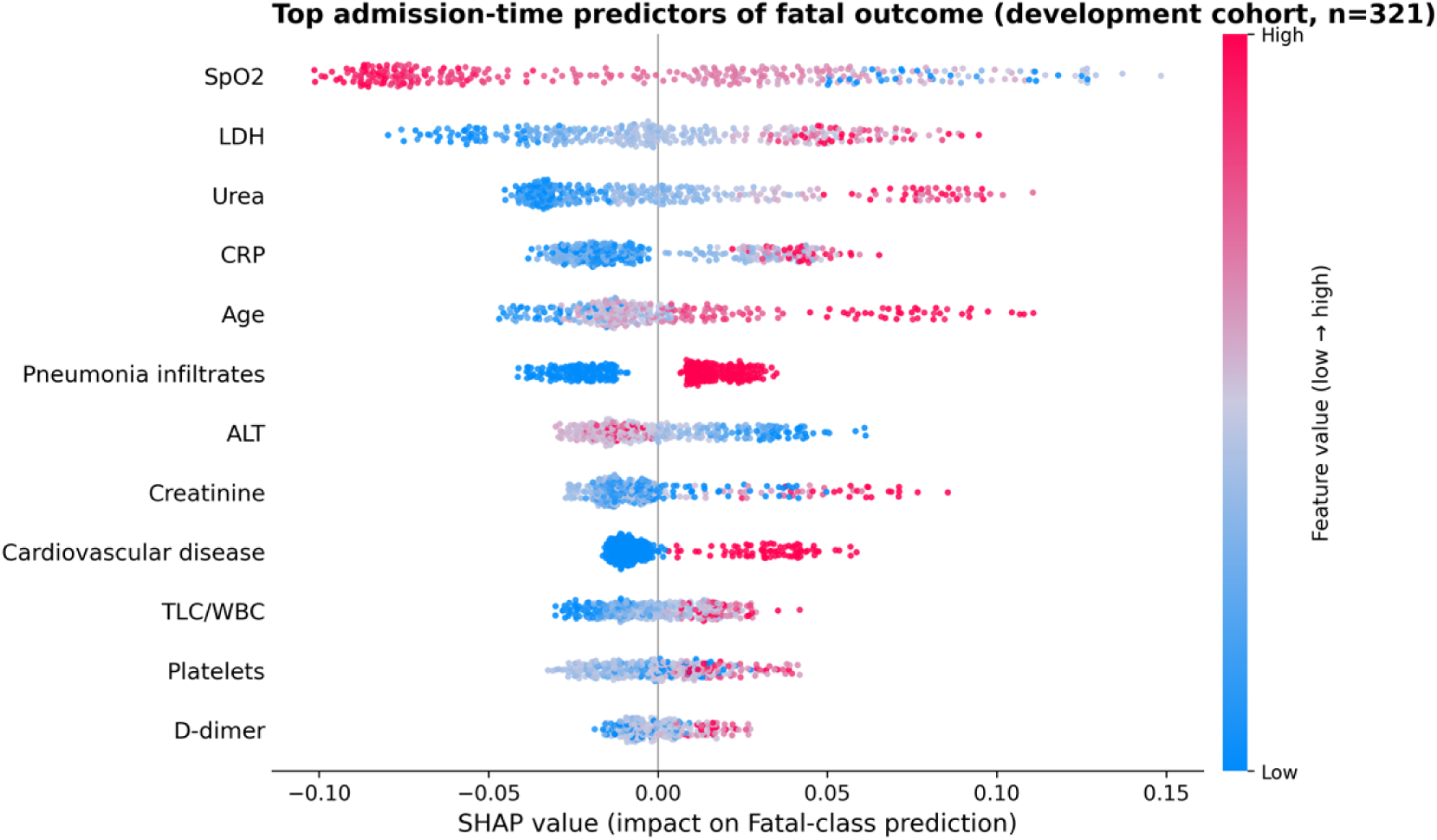
SHAP summary (beeswarm) for the Fatal class in the development cohort (n = 321). Low SpO₂, high LDH, high urea and pneumonia infiltrates drive Fatal-class predictions; colour encodes the feature value (low to high). Features are ordered top to bottom by mean |SHAP| (top 12 of the 20 model features shown).

**Table 3.** Per-class performance of the primary 20-feature model (pooled 10-fold out-of-fold predictions). The Fatal class is identified most reliably; Mild is hardest because of overlap with Severe disease on admission-time markers.

| Class | Precision | Recall | F1 | Support |
| --- | --- | --- | --- | --- |
| Mild | 0.469 | 0.370 | 0.414 | 81 |
| Severe | 0.585 | 0.576 | 0.580 | 132 |
| Fatal | 0.622 | 0.731 | 0.672 | 108 |
| **Macro average** | **0.558** | **0.559** | **0.555** | **321** |

### Model interpretation: a parsimonious set of admission-time predictors

Class-specific SHAP showed that low SpO₂, high LDH, high urea, pneumonia infiltrates and older age drove Fatal predictions (Fig 1). Univariate statistics, serial laboratory trajectories and survival analysis converged on the same predictors. Notably, fatal patients showed a raised urea-to-creatinine ratio: urea was markedly elevated while creatinine rose only modestly (median 0.9 to 1.1 mg/dL, p = 0.11), a prerenal pattern most consistent with dehydration and hypoperfusion rather than intrinsic renal injury [15]. Among binary features, only cardiovascular disease, pneumonia infiltrates, ground-glass opacity and cough were significantly associated with severity (the first three are shown in Table 1); fever, dyspnoea and most other comorbidities were not.

The discriminative value of the laboratory markers was concentrated at admission and decayed thereafter. SpO₂ separated severity strongly on Day 1 (p = 1.1×10⁻⁶), weakly on Day 2 (p = 0.02), and not at all by Day 3, supporting the Day-1 prognosis premise. Treatment-pattern analyses are reported as associational only, since treatment intensity is confounded by indication (sicker patients received more agents); these are summarised in the supplement and are not interpreted causally.

### External validation: discrimination and calibration

Applied to the Chinese iCTCF cohort, the 15-feature model retained useful discrimination: binary mortality AUC 0.821, computed in the 719 of 894 patients with a recorded Cured/Deceased outcome, 57 of them deaths, the remaining 175 being recorded as Unknown, together with severe-or-worse AUC 0.746 and three-class one-vs-rest macro AUC 0.718 across all 894, the Fatal class best separated (AUC 0.816). Threshold-dependent performance was weaker than discrimination alone suggests: macro F1 0.50 and balanced accuracy 0.47, with per-class F1 of 0.833 (Mild), 0.303 (Severe) and 0.352 (Fatal), against a development macro F1 of 0.55. The Fatal class was thus the best separated by AUC and among the weakest by F1: the model ranked Chinese patients by risk well but assigned them to classes poorly, because probabilities learned at a 75% severity prevalence are systematically too high for a cohort at 31% (Fig 2). On the Italian CBC-only cohort discrimination fell to AUC 0.620 (95% CI 0.56-0.68), and to 0.595 (0.52-0.67) after Platt recalibration. Recalibration is a monotonic transformation and cannot itself alter discrimination; the two AUCs differ only because the recalibrated estimate was evaluated on the 80% of the cohort held out from the recalibration fit rather than on the full cohort (Table 4).

**Fig 2.**
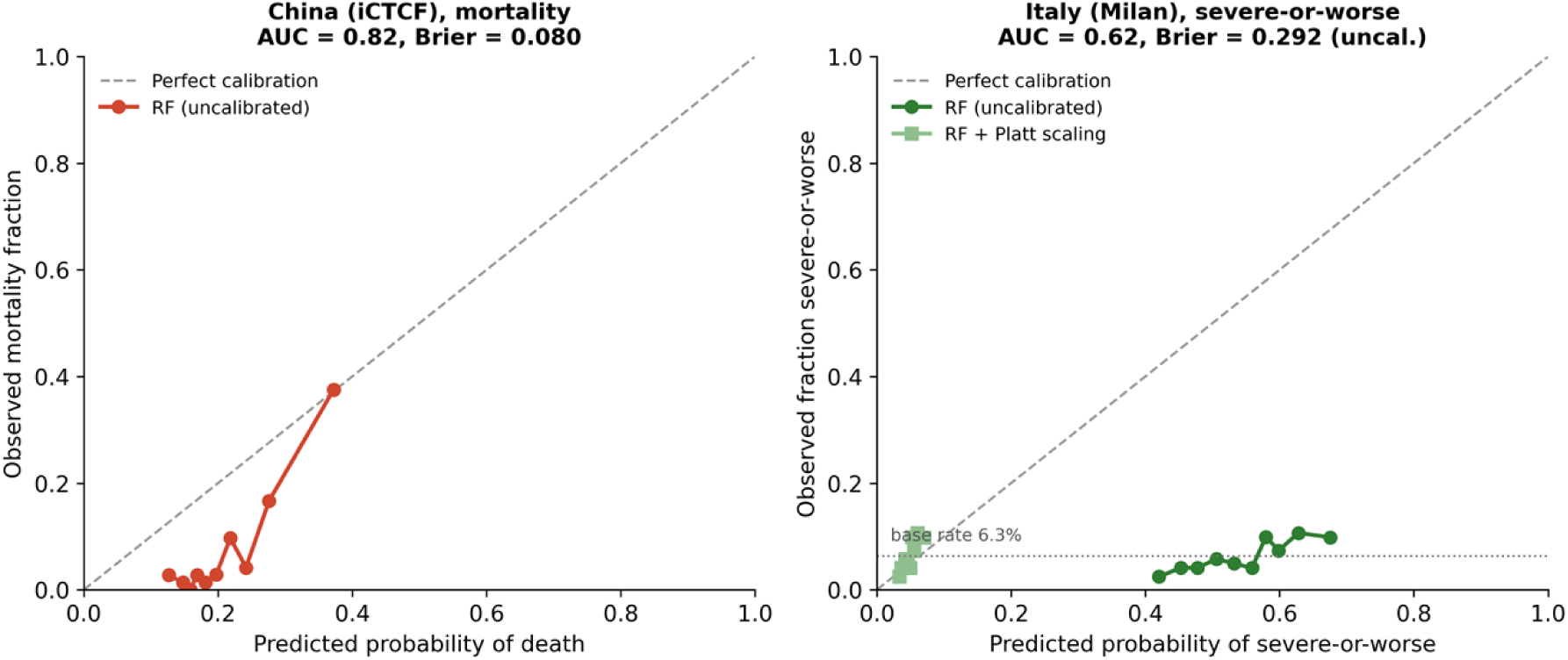
**Calibration curves** on the Chinese cohort (mortality, left) and Italian cohort (severe-or-worse, right). The Chinese mortality predictions lie below the diagonal across the range they span (0.10-0.60), over-predicting death: at a predicted probability of 0.16 the observed mortality is 0.02. The uncalibrated Italian predictions are severely overconfident, with predicted probabilities far above the observed event rate; Platt recalibration to the 6.3% base rate corrects this bias but, because no patient then exceeds the 0.5 decision threshold, eliminates positive predictions.

**Table 4.** External-evaluation summary. Discrimination is lower in the cohort with fewer of the model’s features available; across these two cohorts feature availability is confounded with case-mix and base rate, so the association with feature count is suggestive rather than definitive. Analysis populations differ by endpoint: the Chinese three-class and severe-or-worse endpoints use all 894 patients, the Chinese mortality endpoint the 719 with a recorded outcome (57 deaths), and the Italian severity endpoint all 1,218.

| Cohort | n | Severity prevalence | Features /15 | Severe-or-worse AUC | Mortality AUC |
| --- | --- | --- | --- | --- | --- |
| Pakistan (development) | 321 | 75% | 15 | internal | n/a |
| China (iCTCF, Wuhan) | 894 | 31% | 15 | 0.746 | 0.821 (n = 719) |
| Italy (Milan, CBC) | 1,218 | 6.3% | 5 | 0.620 (0.56-0.68) | n/a (3 deaths) |

Severity prevalence fell sharply across the three cohorts (75% in Pakistan, 31% in China, 6.3% in Italy). As the base rate dropped, the uncalibrated model increasingly over-predicted severe disease, labelling 66% (809 of 1,218) of Italian patients as severe (61 of 77 true positives caught at the cost of 748 false positives). Platt recalibration corrected this bias, but because no patient then exceeded the 0.5 decision threshold under the rescaled probabilities, it eliminated positive predictions entirely (F1 = 0, balanced accuracy 0.50). Recalibration restored the probability scale but could not create discrimination the features did not support (Fig 2).

### Cross-cohort interpretability: stability of model reasoning

The model’s reasoning was highly preserved in China but degraded in Italy (Fig 3, Fig 4). The Spearman correlation of feature importance against the Pakistani baseline was ρ = 0.91 for China and ρ = 0.76 for Italy. In both Pakistan and China the top tier of predictors was the same biochemical cluster (urea, CRP, LDH). In Italy this tier was disrupted, but unevenly. LDH collapsed, falling from the 3rd to the 12th most important feature (mean |SHAP| 0.046 to 0.010, a 77% fall); AST fell by 55% and ALT by 39%, while D-dimer lost only 18% of its magnitude and rose from 9th to 7th, and urea and CRP held both rank and magnitude. This unevenness is a property of the imputation rather than a finding about the model: each of the ten features Italy does not measure was filled by an imputer conditioned on the five it does, leaving them with between 1% and 49% of the spread they have in Pakistan and with no information beyond the complete blood count. Mean |SHAP| is therefore not a safe index of what the model still relies on here; a feature can keep its apparent importance while contributing nothing that was actually observed. Importance redistributed onto the five features Italy actually measures: age, sex, WBC, haemoglobin and platelets (age rose to 1st, WBC to 4th); the combined SHAP mass on these features rose from 22.5% in Pakistan and 21.3% in China to 36.7% in Italy (Fig 3; ranks in Table 5). The loss of discrimination thus coincided with the loss of the model’s biochemical predictors, not the change in population.

**Fig 3.**
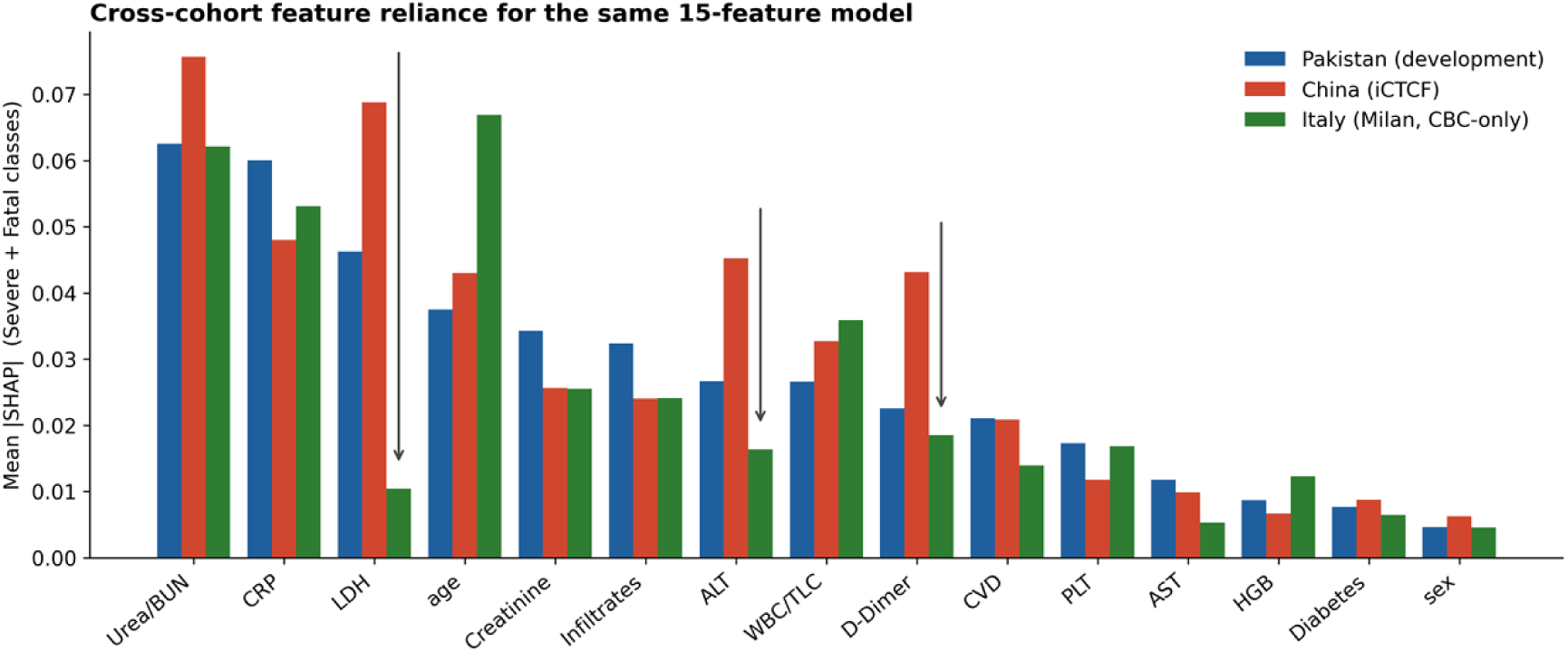
**Mean** |SHAP| (Severe + Fatal classes) per feature for the same 15-feature model applied to each cohort. Biochemical predictors retain weight in China. In Italy the fall is largest for LDH (−77%), then AST (−55%) and ALT (−39%); D-dimer falls only 18%, and urea and CRP retain high apparent importance (0.062 and 0.053) even though they, like the rest of the biochemical panel, are imputed from training-set priors and carry no measured information. Importance also redistributes onto the available CBC features and age.

**Fig 4.**
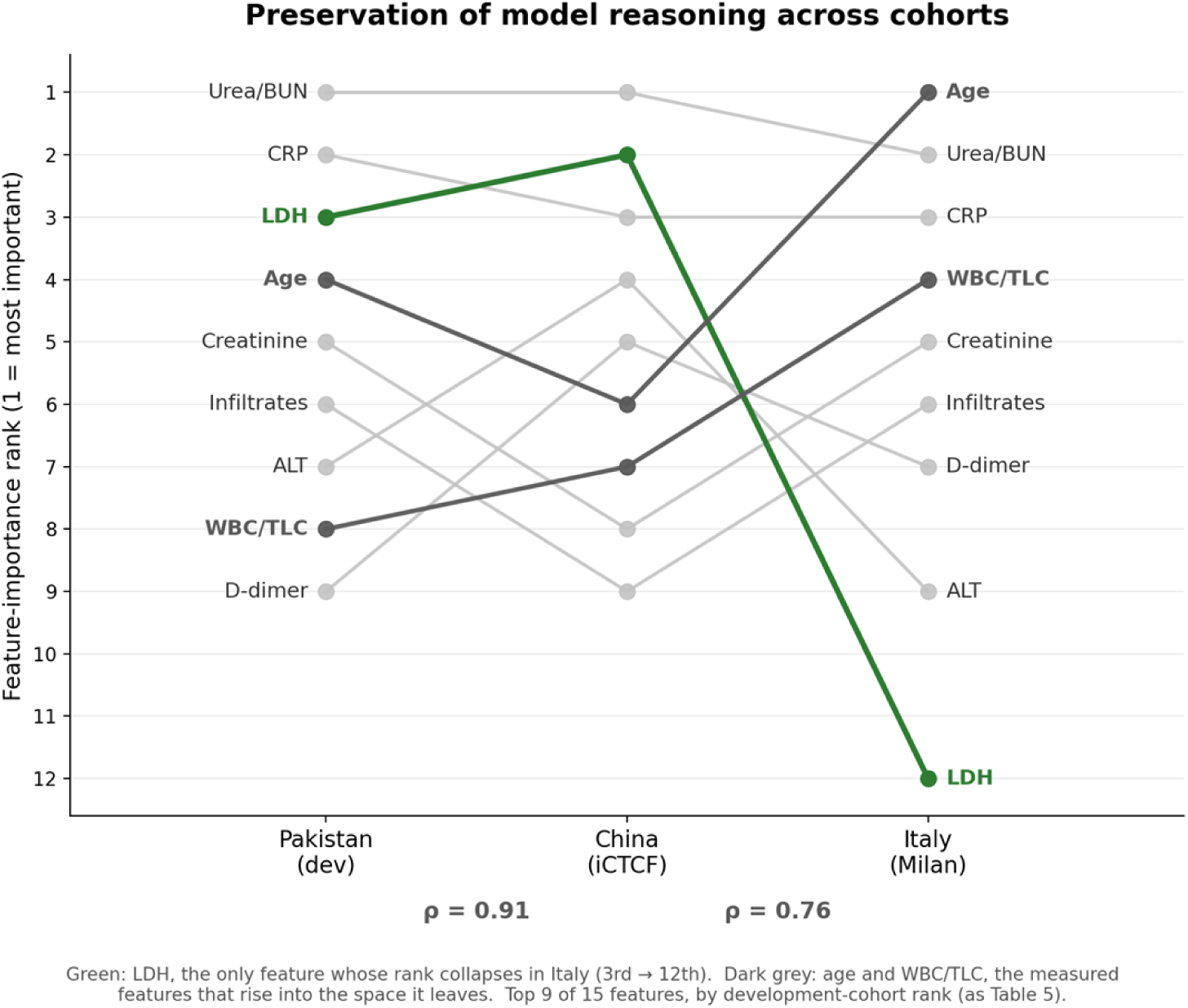
**Feature-importance rank shift** across cohorts. Ranking is preserved from Pakistan to China (Spearman ρ = 0.91) and degraded from Pakistan to Italy (ρ = 0.76). Of the ten features Italy does not measure, only LDH shifts sharply (green: 3rd to 12th); the rest hold or drift, and D-dimer rises from 9th to 7th. The two measured features age and WBC/TLC rise to 1st and 4th to fill the space. Rank stability among the imputed features reflects the imputation model, not a predictor that transferred. Top 9 of 15 features, by development-cohort rank (as Table 5).

**Table 5.** SHAP-based feature-importance rank (Severe + Fatal classes) by cohort. Biochemical predictors retain rank in China. In Italy all ten of them are imputed from training-set priors, yet only LDH (in bold) drops sharply; the others hold rank while carrying no measured information. Top 9 of 15 features, by development-cohort rank.

| Feature | Pakistan rank | China rank | Italy rank |
| --- | --- | --- | --- |
| Urea / BUN | 1 | 1 | 2 |
| CRP | 2 | 3 | 3 |
| **LDH** | 3 | 2 | **12** |
| Age | 4 | 6 | 1 |
| Creatinine | 5 | 8 | 5 |
| Infiltrates | 6 | 9 | 6 |
| ALT | 7 | 4 | 9 |
| WBC / TLC | 8 | 7 | 4 |
| D-dimer | 9 | 5 | 7 |

## Discussion

We developed and externally evaluated an interpretable Day-1 machine-learning model for COVID-19 severity prognosis. Trained on 321 hospitalised PCR-confirmed patients at a Pakistani tertiary care hospital using only admission-time variables, the model identified fatal outcome most reliably (Fatal-class F1 0.67, recall 0.73, within the three-class model) and retained useful mortality discrimination on external testing (mortality AUC 0.82 in a Chinese cohort). These two figures are not directly comparable: one is a threshold-dependent F1 on a three-class endpoint, the other a threshold-free AUC on binary mortality. The model exposes its reasoning at the level of individual features through SHAP and relies on a parsimonious set of admission-time markers (oxygen saturation, lactate dehydrogenase, urea, C-reactive protein, age and radiographic pneumonia). Because these tests require a moderate laboratory panel, the model is best suited to hospitals where that panel is available rather than to the most resource-limited frontline settings, a distinction we return to below.

The features driving the model align with established disease biology rather than appearing arbitrary. SHAP identified a coherent set of admission-time abnormalities spanning three biological domains: low oxygen saturation with radiographic pneumonia, raised inflammatory markers (LDH and CRP), and a raised urea-to-creatinine ratio in fatal cases, with urea markedly elevated and creatinine raised only modestly (p = 0.11). This ratio most likely reflects dehydration (for example from high-grade fever and vomiting) and hypoperfusion rather than intrinsic renal injury; at this sample size the non-significant creatinine rise reflects limited statistical power rather than a confirmed absence of renal involvement. This concordance with the established prognostic value of LDH, CRP and D-dimer in COVID-19 [15–17] and with compact interpretable mortality models built on related markers [18] indicates that the model reacts to recognised prognostic biology rather than to spurious cohort-specific correlations; it does not by itself establish mechanism. Corticosteroid exposure and volume depletion at admission may also contribute to the urea-to-creatinine pattern, so the prerenal reading is one plausible account among several rather than a demonstrated multi-organ mechanism.

On the Chinese iCTCF cohort the model retained good discrimination (mortality AUC 0.82) and its feature-importance ranking correlated strongly with the Pakistani development cohort (ρ = 0.91), indicating that the model relies on the same predictors and reasons the same way in a geographically and ethnically distinct population. The Italian complete-blood-count cohort defined the boundary of transfer: with only a CBC available, the model’s biochemical predictors had to be imputed from priors and contributed almost no patient-specific information; discrimination fell to a modest level (AUC 0.62). Their apparent SHAP importance did not fall uniformly (LDH collapsed while urea and CRP held rank), but that stability is a property of the imputation, not evidence that these markers still contributed. The loss of discrimination coincided with the loss of these predictors, not the change in population, a distinction that a conventional AUC-only validation would have missed. The base-rate shift across cohorts (from 75% in Pakistan to 6.3% in Italy) further illustrates that recalibration to a low-prevalence setting cannot substitute for missing predictors [19].

Two practical implications follow. First, transportability to a new setting appears to depend on the availability of the model’s required admission-time tests; demographic and routine haematology inputs alone were insufficient. Any deployment would require prospective evaluation in the target setting, which our retrospective data cannot substitute for. In LMIC settings where these tests are intermittently available, prioritising their availability is a higher-yield intervention than substituting a CBC-only proxy model. Second, predicted probabilities must be recalibrated to local prevalence, recognising that recalibration cannot substitute for missing predictors. More broadly, our results argue for reporting external validation at the level of model reasoning (for example, with cross-cohort SHAP) alongside discrimination, since the two failure modes (population mismatch versus feature unavailability) have different implications for deployment and revision [8,20].

### Comparison with published work

Our development macro F1 (0.55) is modest beside binary models reporting AUC above 0.85, but it addresses a harder three-class task in a small single-centre sample, prevents leakage (MICE within folds, admission-only features), and is externally validated, a combination rarely reported together and frequently flagged as missing in critical appraisals [3,4,21]. Prospectively validated rapid-triage tools such as the Oxford CURIAL framework [22] illustrate a methodologically rigorous version of this work in a high-income setting. The Pakistani provenance of the development data also places our model in a sparsely populated region of the prediction-model literature, where South Asian COVID-19 cohorts have to date been chiefly addressed by Indian studies [23] and by validation work from other middle-income settings [24,25]. The CBC-only Italian result (AUC about 0.62) is consistent with the literature on out-of-distribution transfer of CBC-based severity models, which perform substantially better when trained and tested on the same dataset [12]. Reporting cross-cohort SHAP rank correlation alongside AUC is, to our knowledge, infrequently done in this clinical literature; we suggest it as a useful complementary descriptor for interpretable-model external-validation studies, while noting that it lacks an established null distribution.

### Limitations

The development sample is small (n = 321, single-centre), which limits the precision of internal performance estimates and may make our cross-validated metrics optimistic relative to true out-of-sample performance [14]. In this context, the Chinese iCTCF mortality AUC of 0.82 is arguably a more honest estimate of the model’s real-world discrimination than the development F1. The external cohorts lack SpO₂ and imaging, so the validated 15-feature variant differs from the full 20-feature model; the external analyses therefore test the transportability of the model’s biochemical core rather than the complete model, and SpO₂, the strongest single predictor in development, could not itself be evaluated externally. Outcome definitions differ across cohorts: the iCTCF “critically ill” category is an imperfect proxy for Fatal, as some critically ill patients survived, and the recorded Cured/Deceased endpoint is the only fully comparable outcome. The Italian cohort did not permit mortality analysis, and its Platt recalibration, although evaluated on a held-out subset, was fitted within the same cohort. Treatment associations are confounded by indication and are not causal. The ρ-SHAP metric we use as a complementary measure of transportability lacks an established null distribution; we present it as descriptive rather than as a hypothesis-test statistic.

Two priorities follow for future work. First, a matched local replication cohort that includes SpO₂ and imaging, together with an additional external cohort with high feature overlap, would allow the complete model to be validated; in particular, validation in culturally and clinically similar South Asian settings would provide a more stringent and arguably fairer test of transportability than the geographically and ethnically distant cohorts examined here. Second, because the admission-time signature the model relies on (hypoxaemia, systemic inflammation and prerenal stress) is not specific to COVID-19, extending and revalidating the approach on other causes of viral and bacterial pneumonia could broaden its relevance as a general resource-limited pneumonia triage tool.

### Conclusion

An interpretable Day-1 machine-learning model identifies high-risk COVID-19 patients in a Pakistani hospital cohort, most reliably those at risk of death, while providing full transparency into the features driving each prediction. The model transfers to a Chinese cohort when its required predictors are available, and identifies a biologically coherent admission-time signature (hypoxaemia, hyperinflammation and prerenal stress) that converges with established markers of severe COVID-19. Across the two external cohorts examined, transferability tracked the availability of the model’s laboratory predictors more closely than either geographic or population distance did, although with only two cohorts, these factors cannot be fully separated. For deployment in hospitals with intermittent laboratory access, prioritising the measurement of these admission-time predictors is the highest-yield path to realising the model’s prognostic value.

## Materials and Methods

### Ethics statement

The study was approved by the Institutional Review Board of Services Institute of Medical Sciences (SIMS), Lahore (IRB/2020/656/SIMS) and the Ethics Review Committee of Lahore University of Management Sciences (LUMS-IRB/05/07/2020SM). The requirement for individual informed consent was waived for this de-identified retrospective analysis in accordance with institutional policy and the Declaration of Helsinki. The Chinese iCTCF and Italian San Raffaele cohorts are publicly available, de-identified datasets used under their respective open-access licences. The study is reported in accordance with the STROBE statement for observational studies [13] and the TRIPOD+AI guidance for prediction-model studies [8].

### Study design and development cohort

We conducted a retrospective cohort study of 321 hospitalised, PCR-confirmed COVID-19 patients admitted to Services Institute of Medical Sciences (SIMS), a tertiary care teaching hospital in Lahore, Pakistan, between 19/04/2020 and 23/03/2021. Data was collected on between 10/5/2020 to 23/8/2021 participants during their admission. The outcome was a three-class ordinal severity label defined from in-hospital course: Mild (not mechanically ventilated and survived), Severe (mechanically ventilated and survived), and Fatal (died in hospital). Ventilation was identified from recorded ventilation dates or days-on-ventilator fields; mortality was taken from the merged final-outcome field, with patients lacking a recorded death assumed to have survived. No patients in this cohort were transferred to another facility or left against medical advice, and patients recorded as discharged were documented as being in good health at discharge, with no annotation to the contrary; the assumption of survival for records without a recorded death was therefore well supported, although rare out-of-hospital deaths after discharge cannot be entirely excluded. Only variables available within 24 hours of admission were eligible as predictors, to avoid leakage from post-admission events and treatments. The Mild and Severe classes were separated by receipt of mechanical ventilation, which is a post-admission clinical decision influenced by bedside judgement, ventilator availability and local protocol as well as by patient physiology; death overrode this assignment, so that a ventilated patient who died was classed Fatal rather than Severe. Of the 211 ventilated patients, 132 survived (Severe) and 79 died, while 29 of the 108 Fatal patients were never ventilated. The Severe label therefore partly encodes a treatment decision rather than a purely physiological state.

### Candidate predictors and data preparation

From 2,278 raw variables in the electronic record extract, clinical knowledge of COVID-19 prognosis was used to define 32 candidate features spanning demographics (age, sex), 11 Day-1 laboratory values (SpO₂, total leucocyte count [TLC], CRP, haemoglobin, creatinine, platelets, LDH, urea, D-dimer, ALT, AST), and 19 binary symptom, comorbidity and imaging flags derived by harmonising free-text records. Radiographic pneumonia was captured through admission chest-imaging flags for infiltrates and ground-glass opacity, which served as the Day-1 imaging proxy for COVID-19 pneumonia. Implausible laboratory values were screened against pre-specified physiological ranges and either corrected or set to missing, with all changes logged for auditability.

### Missing-data handling

Missing values were imputed using multiple imputation by chained equations (MICE), implemented as an iterative imputer with a Bayesian-ridge estimator (20 iterations), with post-imputation clipping to plausible ranges and rounding of binary features [26]. To prevent optimistic bias, the imputer was fitted on the training partition of each cross-validation fold and applied to the held-out partition; the outcome was supplied as an auxiliary variable during fitting only and removed before modelling, in accordance with current best-practice recommendations for prediction-model studies [8,19].

### Model development and selection

A random-forest classifier [27] was chosen for its robustness to mixed-type predictors, its lack of distributional assumptions, and its native interpretability through SHAP. Candidate feature subsets (top 7, 10, 15, 20 by permutation importance, and all 32) were compared by 10-fold stratified cross-validation, optimising macro F1. Permutation importance was computed within the training partition of each cross-validation fold, so that no held-out data contributed to the ranking used for feature selection; feature selection was therefore nested inside the resampling rather than performed once on the full dataset. The 20-feature subset performed equivalently to the full feature set and was retained for parsimony. Three algorithms (random forest, gradient boosting, XGBoost) were tuned by inner grid-search cross-validation; because tuning did not improve out-of-fold performance at this sample size, the default-hyperparameter random forest was retained as the primary model, a phenomenon consistent with the well-described difficulty of stable hyperparameter selection in small clinical cohorts [14]. Class weighting was used throughout to address imbalance. Performance was reported as macro F1, balanced accuracy, accuracy, and per-class precision, recall and F1, all from 10-fold stratified cross-validation. With 108 fatal events and up to 20 candidate predictors, the events-per-variable ratio is approximately 5, which is below conventional minima for prediction-model development. The model carried forward to external evaluation was a 15-feature variant re-fitted with the tuned random-forest hyperparameters (see External-validation cohorts); it is therefore not identical to the primary 20-feature, default-hyperparameter model whose internal metrics are reported above. The tuned hyperparameters were used for the external variant because the reduced feature set was re-fitted as a separate model rather than as a subset of the primary one; readers should note that the internal and external estimates are consequently not from the same fitted model, and the internal metrics should not be read as a baseline against which the external figures measure a change.

### Interpretation and triangulation

Model interpretability was assessed with SHAP using TreeExplainer [5,10], reporting global feature importance, class-specific beeswarm summaries and dependence plots. To triangulate the model’s findings against simpler analyses, we compared its feature ranking against univariate statistics (χ² for binary features, Kruskal-Wallis for continuous features), serial Day 1-5 laboratory trajectories, and treatment-pattern analysis (multivariable-adjusted odds ratios). Because these analyses draw on the same patients and the same pre-selected features, their convergence guards mainly against coding or modelling artefacts rather than providing independent confirmation of generalisable signal.

### External-validation cohorts

Transportability was tested against two independent, publicly available cohorts. The Chinese iCTCF (HUST-19) cohort (Tongji Hospital, Wuhan) contributed 894 PCR-confirmed patients and recorded all 15 transferable features, 13 of them in more than half of patients [11]. The Italian cohort (San Raffaele Hospital, Milan) contributed 1,218 patients (first-observation snapshots) and was complete-blood-count based, recording only 5 of the model features (age, sex, WBC, haemoglobin, platelets) [12]. Five development features absent from both external cohorts (SpO₂, ground-glass opacity, cough, shortness of breath, smoking history) were dropped, and a 15-feature variant of the model was re-fitted on the Pakistani data with the tuned random-forest hyperparameters. Laboratory units were harmonised to the development scale (haemoglobin g/L divided by 10 to g/dL; creatinine μmol/L divided by 88.4 to mg/dL; urea mmol/L multiplied by 6.006 to blood urea nitrogen mg/dL). Features absent from a cohort were imputed with an iterative imputer; for the Italian cohort the imputer was fitted on the Pakistani training data, so its 10 missing features were filled from training-set priors. The iCTCF morbidity category was mapped to the three-class scheme for all 894 patients, and the recorded Cured/Deceased field provided a binary mortality endpoint in the 719 patients for whom it was populated (57 deaths); it is recorded as Unknown for the remaining 175, who are therefore excluded from the mortality analysis only. The Italian endpoint was binary severity; mortality was not analysable owing to only 3 deaths in the first-observation subset.

A third candidate cohort (a 5,644-patient Brazilian dataset) was screened but excluded because its public release provides only z-score-standardised laboratory values and quantile-binned age, precluding the real-unit harmonisation the model requires.

### Cross-cohort interpretability and statistical analysis

To test whether the model relies on the same predictors across settings, TreeExplainer SHAP values were computed for the single 15-feature model applied to each cohort’s feature matrix. Mean absolute SHAP values (Severe + Fatal classes) were ranked per cohort, and the Spearman rank correlation against the Pakistani baseline ranking (ρ-SHAP) quantified the stability of the model’s reasoning. Discrimination was summarised with AUC (one-vs-rest for the three-class endpoint), and uncertainty for the Italian cohort by 1,000-sample bootstrap with Platt recalibration fitted on a 20% stratified subset and evaluated on the remainder. Analyses used Python 3.12 with scikit-learn (v1.3), SHAP (v0.44) and lifelines (v0.27).

## Acknowledgements

The authors thank the clinical and records staff of SIMS, Lahore, for the original data collection during the COVID-19 pandemic, and the iCTCF (HUST-19) and San Raffaele groups for the public release of validation datasets that made this work possible.

## Author contributions

Conceptualization: S.M., M.A.Z. Data curation: S.A., T.J., E.T. Formal analysis: T.J., S.M., E.T. Investigation: S.M., A.K., N.J. Methodology: T.J., S.M., E.T. Project administration: S.M., M.A.Z. Resources: N.J., S.M. Software: T.J. Supervision: S.M., T.M. Validation: T.J. Visualization: T.J. Writing – original draft: T.J. Writing – review & editing: T.J., S.A., N.J., A.K., E.T., T.M., M.A.Z., S.M.

## Data availability

The de-identified patient-level dataset and all analysis code supporting this study are openly available from Zenodo at https://doi.org/10.5281/zenodo.22734717 (CC BY 4.0). Public release of the de-identified data is permitted under the study’s approval from the Services Institute of Medical Sciences review board (IRB/2020/656/SIMS). The raw source records are not publicly available because they contain patient names, medical record numbers, and dates; requests for variables beyond those released should be directed to the corresponding author. The external validation cohorts are third-party data and were not redistributed: the Chinese iCTCF cohort is available at https://doi.org/10.1038/s41551-020-00633-5 and the Italian cohort at https://doi.org/10.5281/zenodo.4686707.

## Footnotes

1 SpO₂ is the strongest predictor in the 20-feature development model but is not recorded in either external cohort, and so is absent from the 15-feature variant used for external evaluation. The cross-cohort importance rankings (Table 5, Figs 3 and 4) are therefore headed by urea rather than SpO₂; the two rankings are not directly comparable, and the apparent demotion of SpO₂ reflects its removal from the model rather than a change in its prognostic value.

